# Deliver in the right place and get there in time! Healthcare-seeking behaviour for delivery in cases of stillbirths and neonatal deaths in rural Cambodia: a prospective cohort social autopsy study

**DOI:** 10.1101/2025.05.19.25327952

**Authors:** Kaajal Patel, Daly Leng, Sophanou Khut, Sothearith Duong, Chou Ly, Arthur Riedel, Koung Lo, Claudia Turner, Verena I. Carrara

## Abstract

**Introduction:** Perinatal mortality remains high in low-resource settings, with many deaths preventable. While clinical causes of stillbirths and neonatal deaths are often examined, non-medical factors such as care-seeking behaviour and barriers are less well understood. This study uses social autopsy to explore upstream, social factors associated with stillbirths and neonatal deaths in rural Cambodia.

**Methods:** A prospective, population-based observational study over three years (2019–2022) in Preah Vihear province, Cambodia. A social autopsy questionnaire to examine socio-demographic characteristics, health-seeking behaviours, and delays in healthcare-seeking was developed. The Three Delays model was used to summarise barriers faced by pregnant women at the time of delivery. Social autopsy interviews were conducted for all stillbirths and neonatal deaths. Data were analysed descriptively.

**Results:** Social autopsy was completed for 315 out of 404 (78.0%) stillbirths and neonatal deaths. Most mothers (87.3%, 275/315) reached a health facility for delivery. However, 20.4% (56/275) of them bypassed their nearest facility. Among women attending their nearest facility, 64.8% (142/219) delivered there, and of these, 69.0% (98/142) delivered within one hour of arrival. In total, nearly half of all deliveries (49.5%) occurred either at home (28/315), enroute (12/315), or within one hour of arrival at the first facility (116/315). Delays to seeking facility-based care for delivery were common: 65.7% (207/315) of women reported experiencing at least one delay type, most often at home (43.5%) or at the facility (36.7%).

**Conclusion:** Healthcare-seeking behaviour was generally appropriate, but not timely. Deliveries occurring very soon after arrival at health facilities likely limited the quality of care that healthcare workers could provide. Addressing home and facility delays may help to reduce these late-presenting deliveries, as well as reducing non-facility births. To improve the timeliness of facility arrival by pregnant women for delivery, we need to better understand the perspectives of families and healthcare workers.

## Introduction

Perinatal mortality rates remain unacceptably high and deaths are predominantly avoidable.^1,2^ Globally, the decline in mortality rates among neonates has been slower than among children under-five.^1^ Understanding the upstream social causes of death can provide a more comprehensive explanation of why these mostly preventable deaths are persisting. Harnessing this knowledge to implement more tailored and holistic strategies across the continuum of care might help to accelerate progress in perinatal mortality rate reductions.^3,4^

Cause of death data is vital for implementing targeted interventions to reduce preventable fatalities. Although identifying the downstream clinical causes of death remains crucial, non-medical factors such as social and health system contexts are critical for comprehensively understanding the circumstances surrounding death.^5^ Factors such as care-seeking behaviours and the quality of healthcare all play vital roles. By examining the social, behavioural, and health system factors contributing to mortality, we may be able to achieve faster reductions in mortality rates.

The delivery of a baby is a critical time in life. Delivery in a health facility with access to skilled care is well-established public health advice aimed at reducing the risk of death of both the mother and baby. However, despite increasing facility-based deliveries, expected declines in perinatal mortality rates have not followed.^6–9^ Prompt, quality healthcare for facility-based deliveries is imperative for improving health outcomes.^10^ However, improving the availability of quality care at birth will not effectively reduce perinatal mortality unless families know when, where, and how to seek it. Insights into healthcare-seeking behaviours and barriers could improve contextual understanding and shed light on tailored public health interventions that save lives.^11^

Although facility-based deliveries are increasing in low-resource settings, many deliveries still occur in the community. Deliveries that occur at home and on the way to health facilities cannot receive quality care at birth. Even when women do reach facilities to deliver, they may arrive with very little time for staff to provide effective clinical care. Delivery soon after facility arrival means potentially limited quality care at birth, which can have important clinical implications. A facility-based stillbirth attributed to intrapartum hypoxia could be prevented by skilled healthcare workers providing rapid, quality obstetric care but only if the pregnant woman arrives at the health facility with sufficient time for assessment and effective management. For facility-based quality care around delivery, pregnant women need to arrive at the right facility at the right time.

For care-seeking to be both appropriate and timely for facility-based deliveries, it is essential to understand the factors involved. This can be achieved firstly, by knowledge of the care-seeking journey taken by women for delivery. Women may bypass their nearest health facility for delivery and/or they may attend multiple facilities prior to delivery. Thus, knowledge of the birth location alone does not provide a complete picture. By describing facilities attended, care-seeking behaviours, journeys, and referral patterns taken during this critical time can be better understood.

Secondly, by understanding the complexities of the care-seeking process for delivery. The Three Delays model serves as a framework to describe the complex processes involved in healthcare seeking and receipt around the time of delivery. The model has been widely used and adapted in the field of maternal and neonatal health to describe the barriers faced by women, babies, and families in accessing healthcare for delivery.^12,13^ The Three Delays model comprises: 1) delay in the decision to seek care, 2) delay in reaching care, and 3) delay in receiving care. The first delay can be further divided into 1) recognition of the problem, and 2) decision to seek care.^14^

Social autopsy (SA) is a valuable epidemiological tool that can provide a comprehensive understanding of the non-medical factors preceding death.^3,4^ It involves data collection using a verbal questionnaire with the family of the deceased on the circumstances surrounding death. Social autopsy enables reporting of the social and behavioural factors, as well as the healthcare seeking processes and barriers encountered. Social autopsy can be combined with verbal autopsy (VA): VA elucidates the ‘what’ and SA elucidates the ‘why’ of mortality.

The main aim of this study was to describe the healthcare seeking patterns and delays encountered by pregnant women that experienced a stillbirth or neonatal death in a rural Cambodian province using social autopsy. We also aimed to describe the socio-demographic, cultural, and behavioural characteristics of the women.

## Methods

This was a three-year (September 2019 to August 2022), prospective observational study covering the North-Eastern Cambodian province of Preah Vihear. This study was nested within the Saving Babies’ Lives (SBL) implementation research programme.^15^ Social autopsy was conducted on all women that experienced a stillbirth or neonatal death.

Bordering Thailand and Laos, Preah Vihear is a large (13,788km^2^) and predominantly agricultural province with 90.2% of the 254,827 population living in rural areas in 2019.^16^ The province is sparsely populated with a population density of 18 persons per km^2^ compared to the national average of 87 persons per km^2^.^16^ The government health facilities in the province comprised 45 primary facilities and two secondary facilities (one referral and one district hospital). Healthcare is free for those with a PoorID card. There are also a few small private clinics. The neighbouring province, Siem Reap, has two tertiary paediatric hospitals run by non-governmental organisations (NGOs), providing free healthcare.

### Study population cohort

Deaths were identified via a population-based neonatal surveillance system, which is described separately [REF VA pre-print]. Deaths resulting from all deliveries ≥28 weeks gestation (or birth weight ≥1000 grams if gestation unavailable) of women living in and/or delivering in the study area during the study date range were included. Stillbirths were defined as babies born with no signs of life, and split by timing of death in relation to labour: antepartum stillbirth (death before onset of labour), intrapartum stillbirth (death during labour), and stillbirth of unknown timing. Neonatal deaths were defined as babies born with any sign(s) of life and death during the first 28 completed days of life, and were split by timing of postnatal death: early neonatal death (within the first seven completed days of life) and late neonatal deaths (death after the first seven days of life).

### Data collection: Social Autopsy tool

Timing and medical causes of deaths were already classified with VA methodology [REF VA pre-print]. The VA tool was merged with our SA questionnaire, creating a VASA tool. VASA was performed within six months of death for all stillbirths and neonatal deaths by a trained study team using a tablet-based application with a close caregiver of the deceased. Interviewers had been working in the study area for several years and knew the area well.

We developed a comprehensive SA tool, tailored to our context, and refined to reflect preliminary findings. The SA tool covered social or non-medical features prior to stillbirth or neonatal death, as described by interviewees. The SA tool was divided into four main parts and focussed on healthcare seeking behaviours of pregnant women for delivery, prior to stillbirths and neonatal deaths:

1. Characteristics: Clinical, socio-demographic, obstetric history, antenatal care-seeking and behaviour, and cultural (traditional practice use) factors.
2. Health journeys: Health facilities attended for delivery (if any) by pregnant women in labour or experiencing complications.
3. Healthcare seeking process: detail of the healthcare seeking process of pregnant women from home to the first facility they attended.
4. Healthcare seeking delays: home, transport and healthcare delays experienced by pregnant women from home to the first facility attended.

Community health workers (CHWs), who ran the surveillance system to discover deaths, helped organise and coordinate the SA interviews. Interviews mostly took place in the interviewee’s home.

### Data analysis

All analysis was performed using the R software package.^17^ Descriptive statistics were used to describe the socio-demographic, cultural, behavioural, family, and antenatal characteristics of women and families in the social autopsy cohort. Where relevant, the cohort was stratified into four groups based on timing of death: 1) unknown timing of stillbirth, 2) antepartum stillbirth, 3) perinatal death (intrapartum stillbirth or early neonatal death), and 4) late neonatal death. Of note, the use of the term ‘perinatal death’ hereafter includes only intrapartum stillbirths and neonatal deaths within the first seven completed days of life; the term excludes stillbirths that were of unknown timing or antepartum.

### Health journeys

To visualise the number, sequence, and type of facilities visited by women prior to delivery we used Sankey diagrams. We designated locations according to the health system structure (Appendix 1 for detailed definitions). Firstly, within the study area (Preah Vihear province): home, enroute (home to health facility or between health facilities), government sector comprising ‘PrimaryA’ (minor primary facility), ‘PrimaryB’ (normal primary facility), ‘SecondaryA’ (lowest level referral hospital), ‘SecondaryB’ (provincial referral hospital) and private sector. Secondly, outside Preah Vihear province: other province Primary / Secondary / Private / NGO which are based on the previous definitions. Of note all NGO hospitals are tertiary-level hospitals and are located out of province.

### Healthcare seeking process

To detail the context of the Sankey plots, particularly focussed on the initial part from home to the first facility, descriptive statistics were used. For cases that attended a facility, we explored whether pregnant women attended or bypassed their nearest facility and the time from arrival at this facility to delivery.

### Healthcare seeking delays

Descriptive statistics were used to describe the barriers encountered by women during the initial part of their health journey, from home to the first facility they attended for delivery. Delays were assigned based on responses, using an adapted three delays framework, which involves delays at home (delay 1), transport (delay 2), and facility (delay 3) (Figure 1).^18,19^

**Figure 1:**
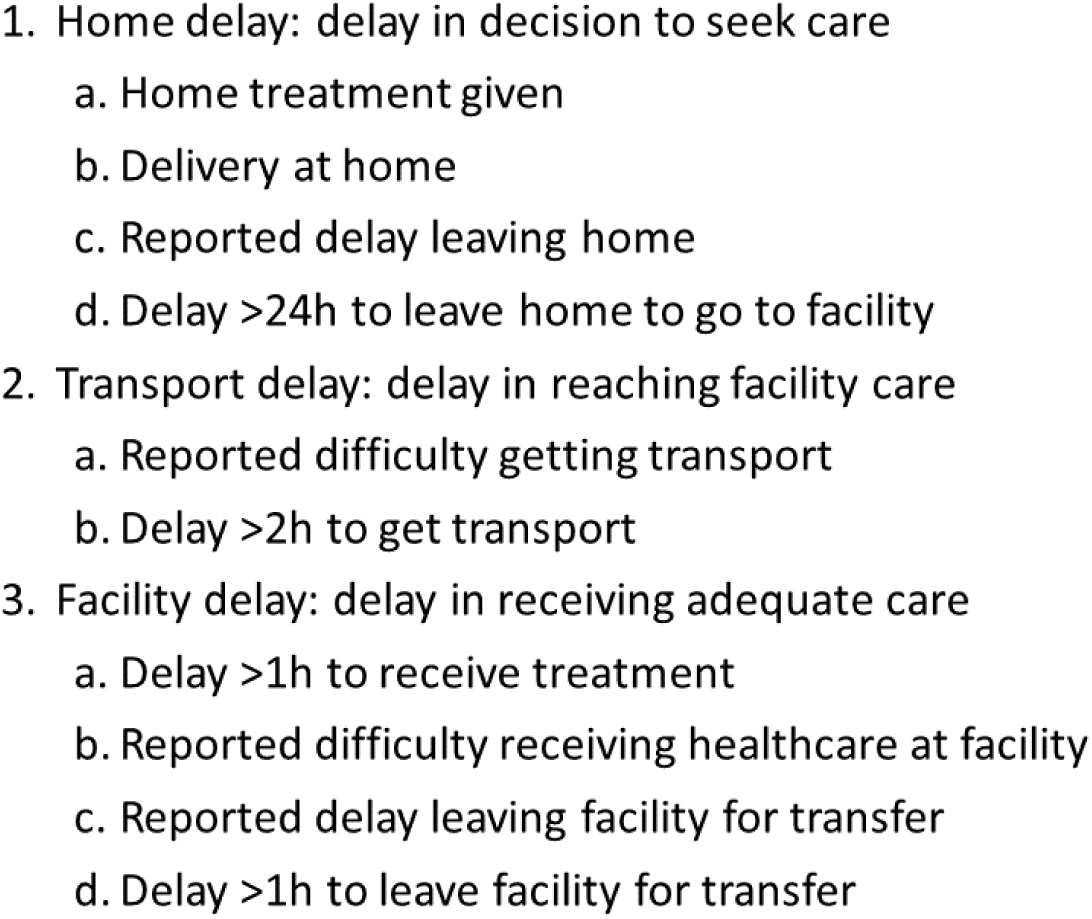
Three delays definitions.

### Patient and Public Involvement

Pregnant women, fathers and families that had experienced a stillbirth or neonatal death were not involved in setting the research question, study design, or SA tool development. We have already disseminated the main results to other key stakeholders including CHWs, healthcare workers, healthcare leaders in the province, and donors. After publication, we will seek an appropriate method of dissemination to share our key results for a non-specialist audience, including the families involved in this study and the public.

### Ethics

The sensitive nature of SA required careful ethical consideration. Social autopsy was conducted after allowing for a mourning period, and confidentiality of cases and of participant views was always maintained. The study was approved by the Cambodian National Ethics Committee for Health Research (NECHR, 283) and the Oxford Tropical Research Ethics Committee (OxTREC, 547–17).

## Results

Social autopsy was conducted on 315 out of 404 (78.0%) stillbirths and neonatal deaths recorded in Preah Vihear province, Cambodia over three years. There were 14 (4.4%) stillbirths of unknown timing, 72 (22.9%) antepartum stillbirths, 203 (64.4%) perinatal deaths (41 intrapartum stillbirths and 162 early neonatal deaths), and 26 (8.3%) late neonatal deaths. Reasons for non-enrollment included inability to find the family, road access issues, and deaths being reported too late (>six months) (Figure 2). The median time from date of death to SA interview was 59.0 days (IQR 34.5-98.5 days). The mother was the main SA respondent for 98.1% (309/315) of cases.

**Figure 2:**
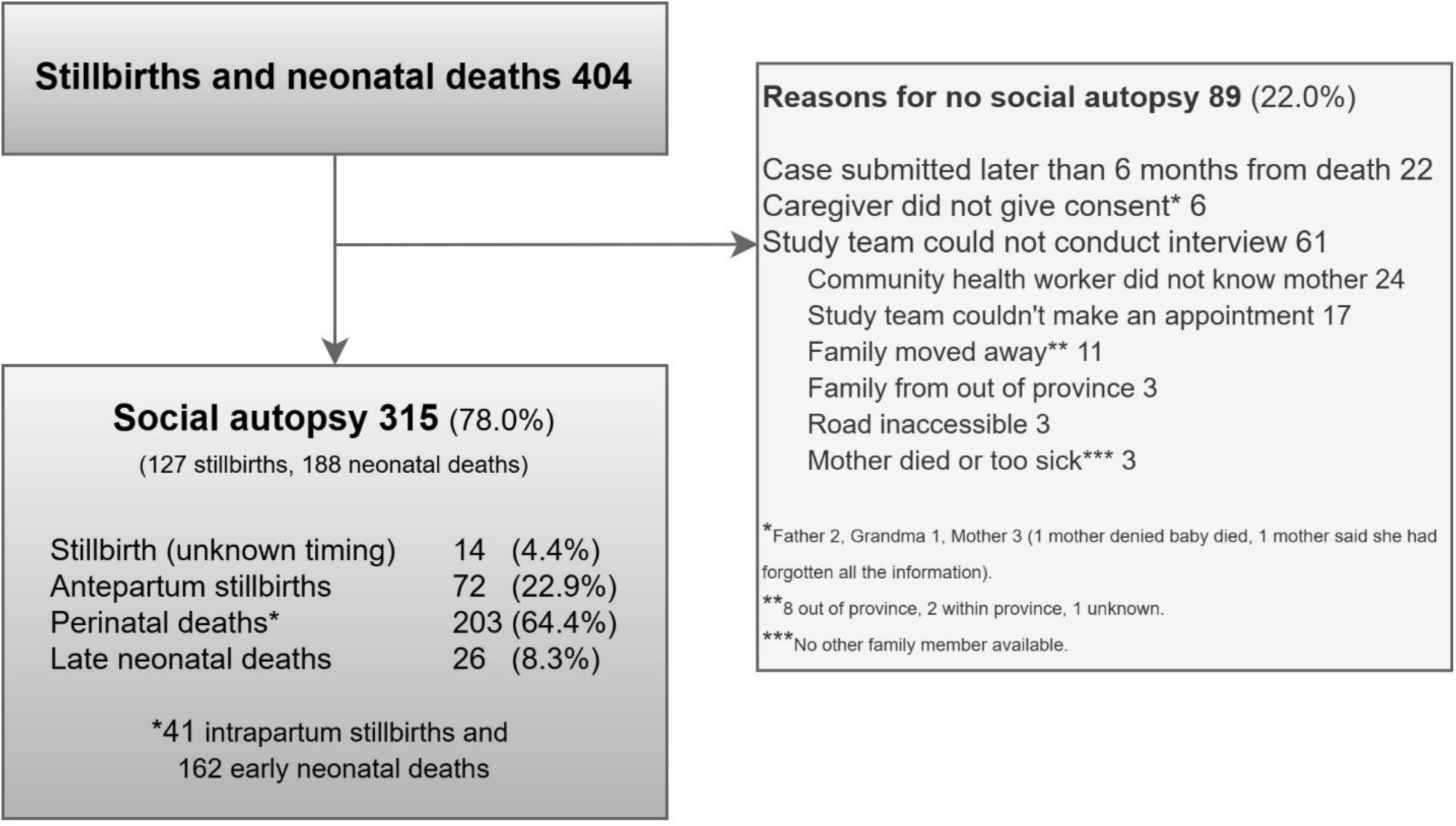
Study flowchart

### Cohort characteristics

### Clinical characteristics

Baseline characteristics of the SA cohort, stratified by timing of death, are shown in Table 1. Of the 315 deaths, 62.5% (197) were male, 53.0% (167) were preterm, the median birth weight was 2.20kg (IQR 1.50, 3.00), and the top causes of death were hypoxia (28.9%), low birth weight and prematurity (23.5%), and infection (12.4%). Of the 315 deliveries, 84.1% (265) occurred in a health facility. Of the 188 live births, 73.4% (138) died in a health facility. There were 276 normal vaginal, 16 vacuum, 2 forceps, 20 caesarean, and 1 postmortem (caesarean done in dead mother by family) deliveries.

**Table 1:** Characteristics of social autopsy cohort.

| Characteristic | Stillbirth (unknown timing)<br>N = 14 | Antepartum stillbirth<br>N = 72 | Perinatal death<br>N = 203 | Late neonatal death<br>N = 26 | Overall<br>N = 315 |
| --- | --- | --- | --- | --- | --- |
| **Gender, n (%)** |  |  |  |  |  |
| Female | 2 (14.3%) | 30 (41.7%) | 73 (36.0%) | 12 (46.2%) | 117 (37.1%) |
| Male | 12 (85.7%) | 41 (56.9%) | 130 (64.0%) | 14 (53.8%) | 197 (62.5%) |
| Unknown | 0 (0.0%) | 1 (1.4%) | 0 (0.0%) | 0 (0.0%) | 1 (0.3%) |
| **Gestation, n (%)** |  |  |  |  |  |
| Preterm | 5 (35.7%) | 39 (54.2%) | 111 (54.7%) | 12 (46.2%) | 167 (53.0%) |
| Term | 9 (64.3%) | 33 (45.8%) | 92 (45.3%) | 14 (53.8%) | 148 (47.0%) |
| **Birth weight (kg), median (IQR)** | 2.90 (1.90, 3.60) | 2.60 (1.90, 3.15) | 2.00 (1.40, 3.00) | 2.45 (1.30, 3.30) | 2.20 (1.50, 3.00) |
| Missing | 4 | 16 | 21 | 2 | 43 |
| **Cause of death, n (%)** |  |  |  |  |  |
| Hypoxic event | 0 (0.0%) | 5 (6.9%) | 85 (41.9%) | 1 (3.8%) | 91 (28.9%) |
| Infection | 0 (0.0%) | 4 (5.6%) | 24 (11.8%) | 11 (42.3%) | 39 (12.4%) |
| Congenital malformations | 1 (7.1%) | 1 (1.4%) | 10 (4.9%) | 6 (23.1%) | 18 (5.7%) |
| Low birth weight and prematurity | 0 (0.0%) | 0 (0.0%) | 67 (33.0%) | 7 (26.9%) | 74 (23.5%) |
| Disorders related to fetal growth | 0 (0.0%) | 1 (1.4%) | 0 (0.0%) | 0 (0.0%) | 1 (0.3%) |
| Other condition | 0 (0.0%) | 1 (1.4%) | 1 (0.5%) | 0 (0.0%) | 2 (0.6%) |
| Unknown | 13 (92.9%) | 60 (83.3%) | 16 (7.9%) | 1 (3.8%) | 90 (28.6%) |
| **Birth location, n (%)** |  |  |  |  |  |
| Home | 1 (7.1%) | 7 (9.7%) | 16 (7.9%) | 4 (15.4%) | 28 (8.9%) |
| Enroute | 2 (14.3%) | 3 (4.2%) | 14 (6.9%) | 3 (11.5%) | 22 (7.0%) |
| PrimaryA | 1 (7.1%) | 1 (1.4%) | 9 (4.4%) | 1 (3.8%) | 12 (3.8%) |
| PrimaryB | 4 (28.6%) | 14 (19.4%) | 76 (37.4%) | 10 (38.5%) | 104 (33.0%) |
| SecondaryA | 0 (0.0%) | 2 (2.8%) | 8 (3.9%) | 1 (3.8%) | 11 (3.5%) |
| SecondaryB | 2 (14.3%) | 31 (43.1%) | 64 (31.5%) | 4 (15.4%) | 101 (32.1%) |
| Private | 0 (0.0%) | 1 (1.4%) | 1 (0.5%) | 0 (0.0%) | 2 (0.6%) |
| Other province Primary | 0 (0.0%) | 1 (1.4%) | 2 (1.0%) | 0 (0.0%) | 3 (1.0%) |
| Other province Secondary | 2 (14.3%) | 2 (2.8%) | 2 (1.0%) | 0 (0.0%) | 6 (1.9%) |
| Other province NGO | 2 (14.3%) | 10 (13.9%) | 11 (5.4%) | 2 (7.7%) | 25 (7.9%) |
| Other province Private | 0 (0.0%) | 0 (0.0%) | 0 (0.0%) | 1 (3.8%) | 1 (0.3%) |

### Social characteristics

The median maternal age was 26 (IQR 20, 32). Most mothers (67.6% (213/315)) and fathers (74.9% (236/315)) worked in agriculture. Most mothers had completed at least primary-level education (84.1% (265/315)); 15.3% (48) reported no education. Nearly half (49.5%) of mothers reported not saving any money during pregnancy.

For 35.2% (111) of all women this was their first delivery (primiparous), and 26.3% (83/315), 33.3% (105/315) and 5.1% (16/315) of women had one, between 2-4 and ≥5 previous deliveries respectively. The median time interval since the last pregnancy was 49 (31-75) months. Of all women in this study, 11.3% (23) and 10.8% (22) had had at least one previous stillbirth or neonatal death respectively.

Only 2.9% (9) pregnant women had no antenatal care; 47.3% attended four or more antenatal appointments. Nearly all pregnant women reported taking iron (95.2%, 300) and folic acid (92.7%, 292) supplements antenatally, although less (72.4%, 228) reported taking deworming treatment. The median number of antenatal ultrasound scans was 2.0 (IQR 2.0, 4.0) with only 19 (6.1%) women not having any scan. Few women used tobacco (7.3% (23/315)) or alcohol (9.5% (30/315)) during pregnancy. During the month prior to delivery 39.7% (125/315) of women did at least one day of manual work, with 18.1% (57/315) of them doing this daily. Three mothers died during the postnatal period.

Many pregnant women reported use of traditional practice (most commonly tea brewed with tree bark), especially postnatally (67.6% (213/315)), compared to antenatally and during labour (33.0% (104/315) and 5.4% (17/315) respectively) (Appendix 2). Postnatally, many mothers also used a hot bed (61.6% (194/315), accompanied by their baby in 18 cases (Appendix 3). The median duration of hot bed use by both mother and baby was 7 days (IQR 5, 7). Amongst 112 neonates that survived more than 12 hours, use of traditional practice was rare (10.7%, 12/112), as was application of any non-medical substance to the umbilical cord (4.5% (5/112)) (Appendix 4).

### Health journeys

The successive health facilities attended (if any) by pregnant women prior to the delivery of a stillbirth or early neonatal death are shown as Sankey diagrams in Figure 3. Health journeys of pregnant women prior to delivery of antepartum stillbirths were complex and prolonged, with multiple facilities visited. For perinatal deaths, health journeys were less complex, with most women delivering at the first facility they attended. Both Sankey pathways demonstrate that with each progressive facility attended, pregnant women generally moved up the health system, to a higher level of care, prior to delivery.

**Figure 3:**
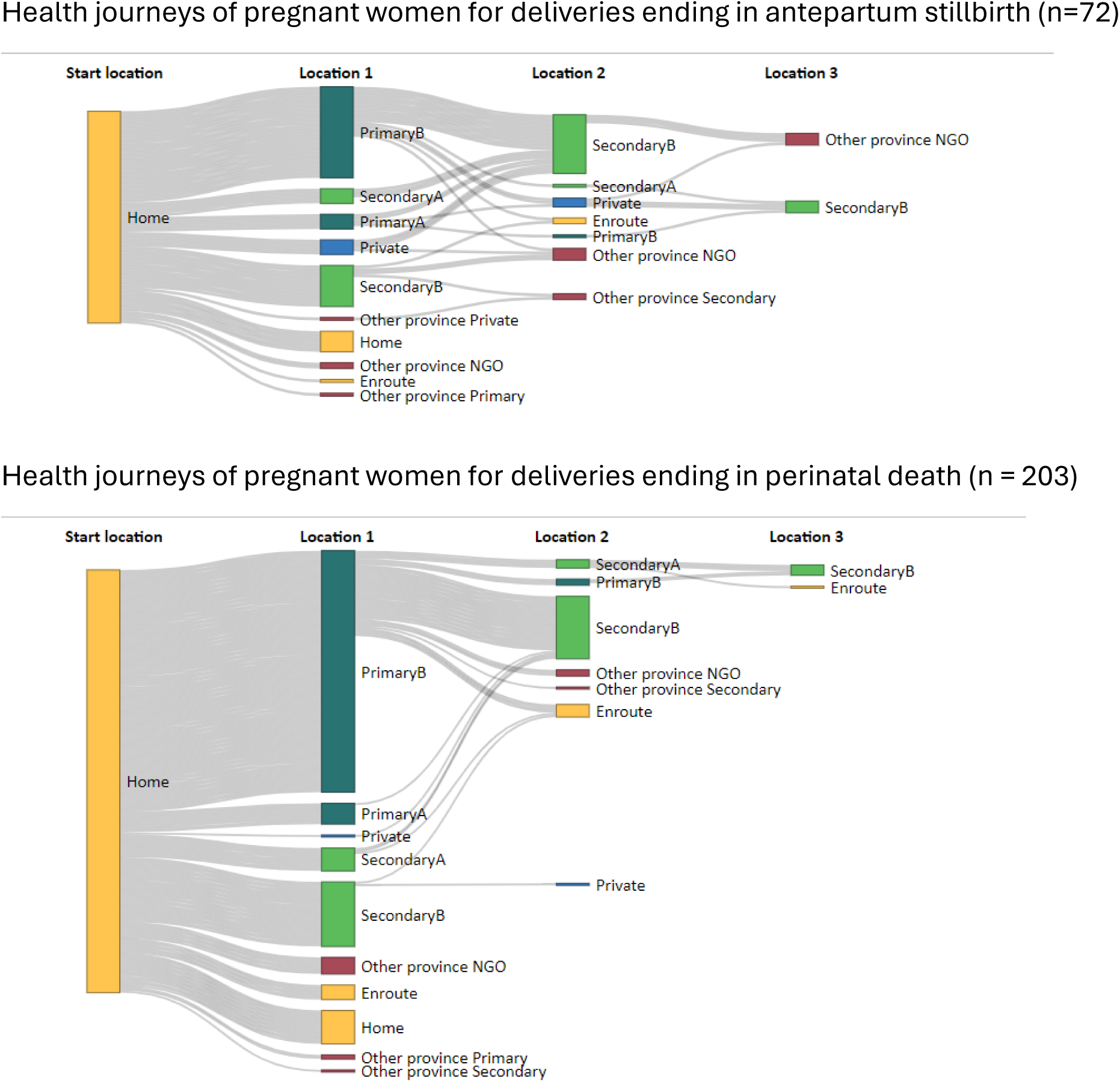
Sankey diagrams show the pathways through the healthcare system taken by women for delivery prior to stillbirth or neonatal death. The pathway starts on the left with all women at home and the pathway ends to the right when the woman delivers. Moving from left to right the diagram shows each subsequent healthcare facility encounter. Thus, the pathways of women that visited several facilities prior to delivery extend further to the right. A home birth is indicated by ‘Home’ for Location 1. The width of the left-to-right flowing grey bars represents the proportion of women travelling to each facility type. The height of the coloured nodes represents the proportion of women presenting to that facility. The colours represents health system levels (yellow: community, dark green: in province primary, light green: in province secondary, blue: in province private, red: out of province facility)

### Healthcare seeking process

Figure 4 provides more detail about the first (left-hand side) part of the Sankey pathways in Figure 3: the movement of pregnant women from home to the first facility they attended prior to delivery of a stillbirth or neonatal death. Out of 315 deaths, 28 delivered at home, 12 delivered enroute from home to a facility, and 275 (87.3%) arrived at a facility for delivery. Of these 275 cases, 79.6% (219/275) attended their nearest facility, which was a government facility within the province in all cases (79.5% primary and 20.5% secondary facility). Amongst the 56 women who bypassed their nearest facility for delivery, 21.6% and 35.7% presented at a primary and secondary facility within the province respectively; and 32.1% sought higher-level healthcare outside of the province. The median time taken to travel from home to the first facility was less for pregnant women who presented at their nearest facility (20 minutes (IQR 10, 30)) compared to those that bypassed it (60 minutes (IQR 28, 110)). Most women presenting at their nearest facility travelled by either motorcycle (54.3%, 75/275) or tractor (34.2%, 75/275). Timing and cause of deaths were similar whether the woman went first to her nearest facility or bypassed it for delivery (Appendix 5).

**Figure 4:**
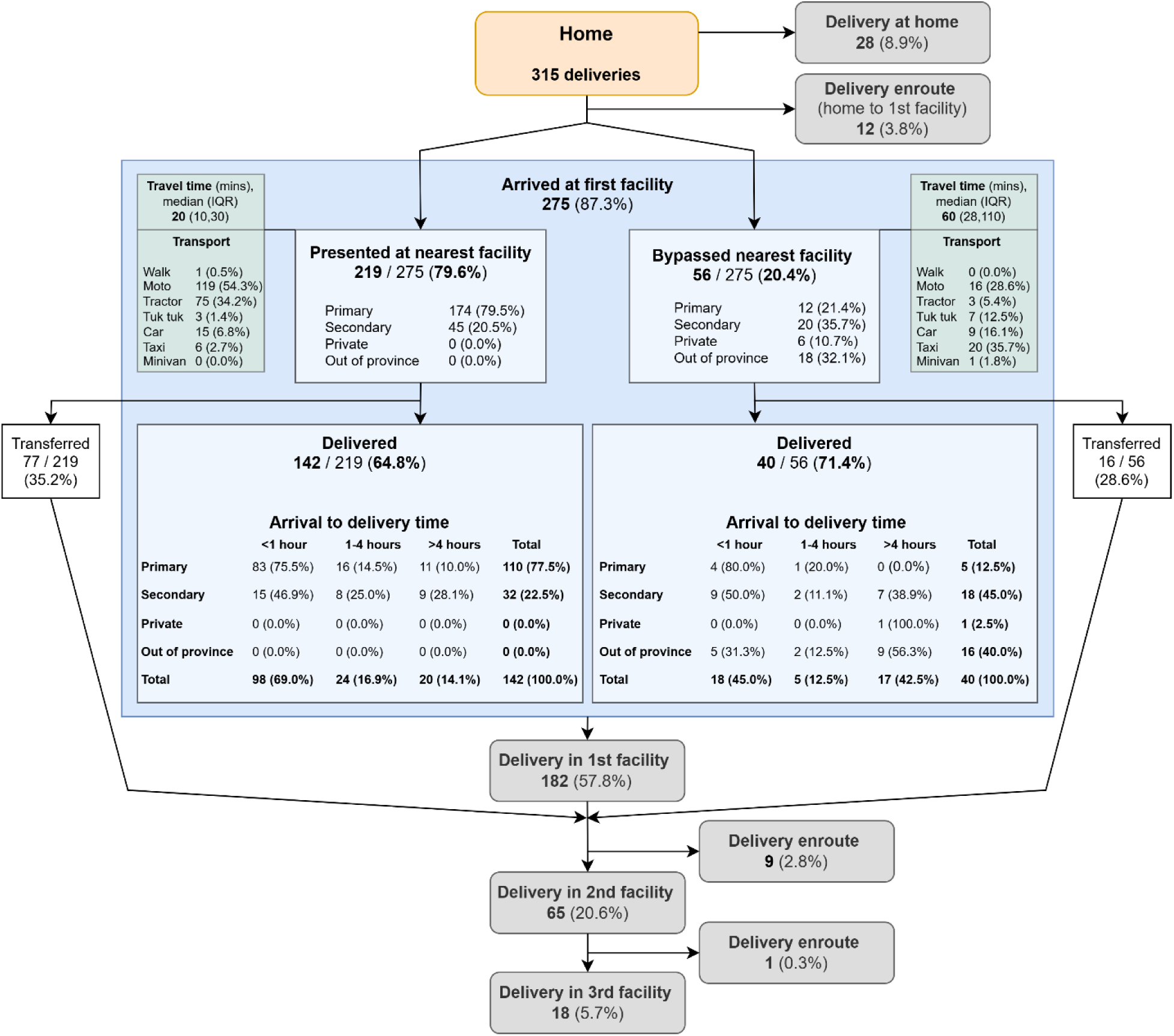
Flow chart of where all 315 deliveries occurred prior to stillbirth or neonatal death. All cases started at home. The delivery location is shown in the grey boxes. All data shown within the large blue box relates to the first facility the women attended from home, and is split by whether that facility was their nearest facility or not (bypassed). If the women delivered at this first facility they attended, the arrival to delivery times by health system level are shown in the tables.

Of the 219 cases that presented at their nearest facility, 64.8% (142) delivered at this same facility, and the remainder (77, 35.2%) were transferred out. Amongst the 142 women that delivered at their nearest facility, 69.0% (98) delivered within one hour of arrival, and this proportion was higher if the nearest facility was primary (75.5%, 83/98). In contrast, amongst women that bypassed their nearest facility and delivered at the first facility they attended, only 45.0% (18/40) delivered within one hour of arrival. Of the 56 cases where the pregnant woman bypassed their nearest facility, a greater proportion, 71.4% (40/56), delivered at that facility.

In total, nearly half of all deliveries (49.5%) occurred either at home (28/315), enroute (12/315), or within one hour of arrival at the first facility (116/315).

### Healthcare seeking delays

Of all 315 deaths included in this study, 65.7% (207) were associated with the pregnant mother experiencing at least one out of the three delay types (home, transport, and facility) prior to delivery (Table 2). Only 4.4% (14/315) reported experiencing all three delays. Perinatal and late neonatal deaths were particularly associated with the mother reporting no delays prior to delivery (36.5%, 74/203 and 38.5%, 10/26 respectively) (Table 2).

**Table 2:** Delays experienced by pregnant women for deliveries ending in stillbirth and neonatal death, by timing of death.

| Type of delay(s)<br>reported per case | Stillbirth<br>(unknown<br>timing)<br>N = 14 | Antepartum<br>stillbirth<br>N = 72 | Perinatal<br>death<br>N = 203 | Late neonatal<br>death<br>N = 26 | Overall<br>N = 315 |
| --- | --- | --- | --- | --- | --- |
| **None** | 4 (28.6%) | 20 (27.8%) | 74 (36.5%) | 10 (38.5%) | 108 (34.3%) |
| **Delay 1** | 3 (21.4%) | 20 (27.8%) | 38 (18.7%) | 8 (30.8%) | 69 (21.9%) |
| **Delay 2** | 0 (0.0%) | 1 (1.4%) | 10 (4.9%) | 3 (11.5%) | 14 (4.4%) |
| **Delay 3** | 3 (21.4%) | 11 (15.3%) | 31 (15.3%) | 2 (7.7%) | 47 (14.9%) |
| **Delay 1+2** | 1 (7.1%) | 7 (9.7%) | 13 (6.4%) | 2 (7.7%) | 23 (7.3%) |
| **Delay 1+3** | 2 (14.3%) | 11 (15.3%) | 17 (8.4%) | 1 (3.8%) | 31 (9.8%) |
| **Delay 2+3** | 1 (7.1%) | 1 (1.4%) | 7 (3.4%) | 0 (0.0%) | 9 (2.9%) |
| **Delay 1+2+3** | 0 (0.0%) | 1 (1.4%) | 13 (6.4%) | 0 (0.0%) | 14 (4.4%) |

Delays at home (delay one) were the commonest reported delay type (43.5%, 137/315), particularly for antepartum stillbirths (53.4%) and late neonatal deaths (57.9%) (Figure 5 and Appendix 6 and 7). Facility delays (delay three) were the second commonest (36.7%, 101/275), and transport delays (delay two) the least common (20.9%, 60/287) (Figure 5). This order was the same regardless of timing of death, except for late neonatal deaths, which had more transport delays than facility delays (Appendix 6 and 7). Home delays were associated with deaths due to infection and facility delays were associated with hypoxia-related deaths (Appendix 8).

**Figure 5:**
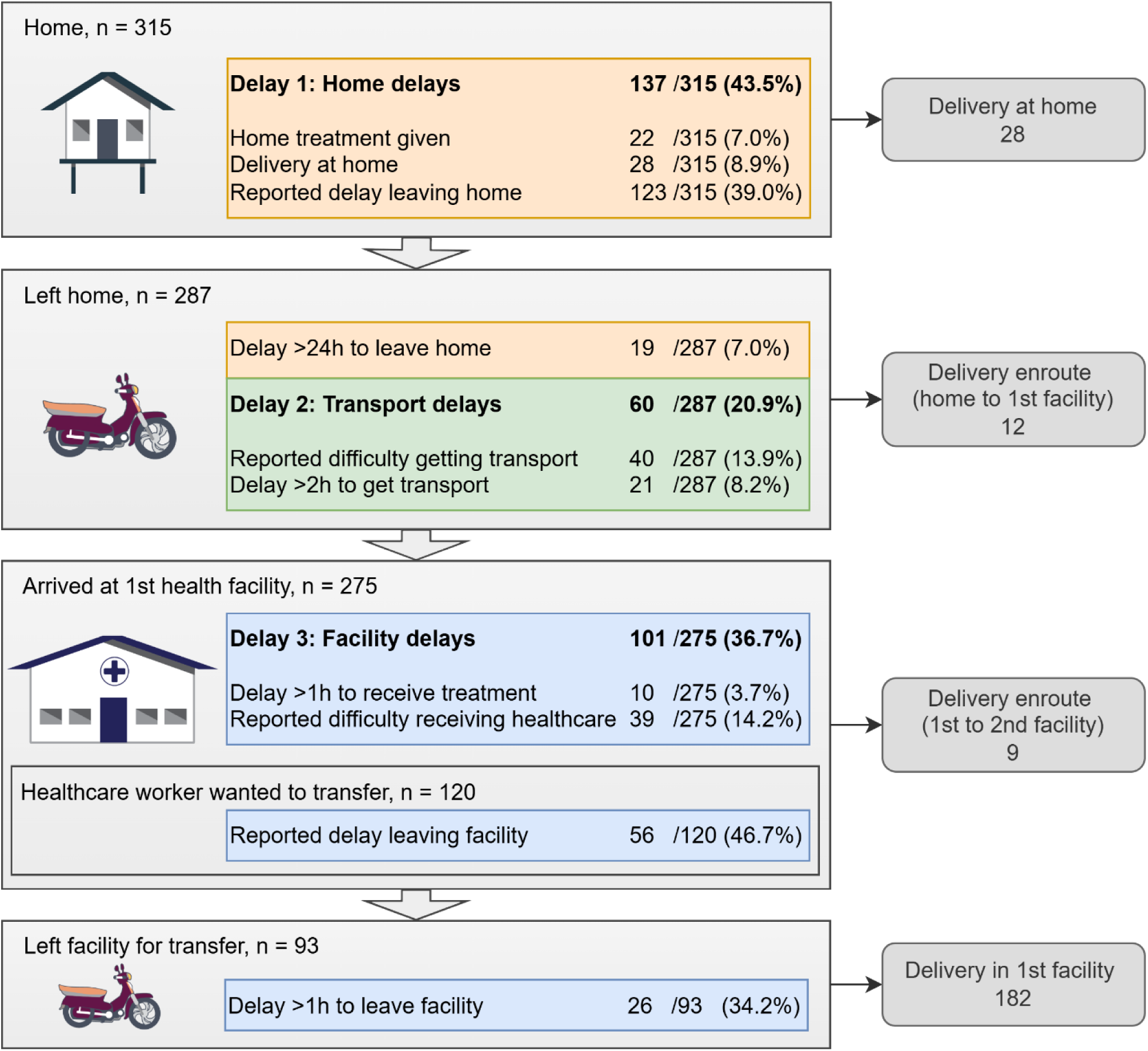
Summary of delays reported by pregnant women for delivery prior to a stillbirth or neonatal death, shown using the Three Delays framework (home, transport, facility). Details for each delay type are shown in Appendix 6 and 7.

A variety of reasons were reported for home delays experienced by pregnant women prior to delivery, including not recognising labour (9.8%), lack of transport and/or driver (10.2%), waiting for a family member to arrive (6.7%), and hoping for an improvement in the mother’s condition (5.7%) (Appendix 9). Reported reasons for facility delays in transferring the pregnant women to a higher level of care included a too rapid delivery (13.3%), lack of transport and/or driver (12.5%), and the mother returning home to prepare her things (5.8%) (Appendix 9).

For pregnant women that delivered at their first and nearest health facility, the proportion of home delays was highest (46.1%) for those that delivered within one hour of arrival and lowest for those that delivered after four hours of arrival (Figure 6). When the arrival to delivery time was more than one hour, facility delays was the major delay type.

**Figure 6:**
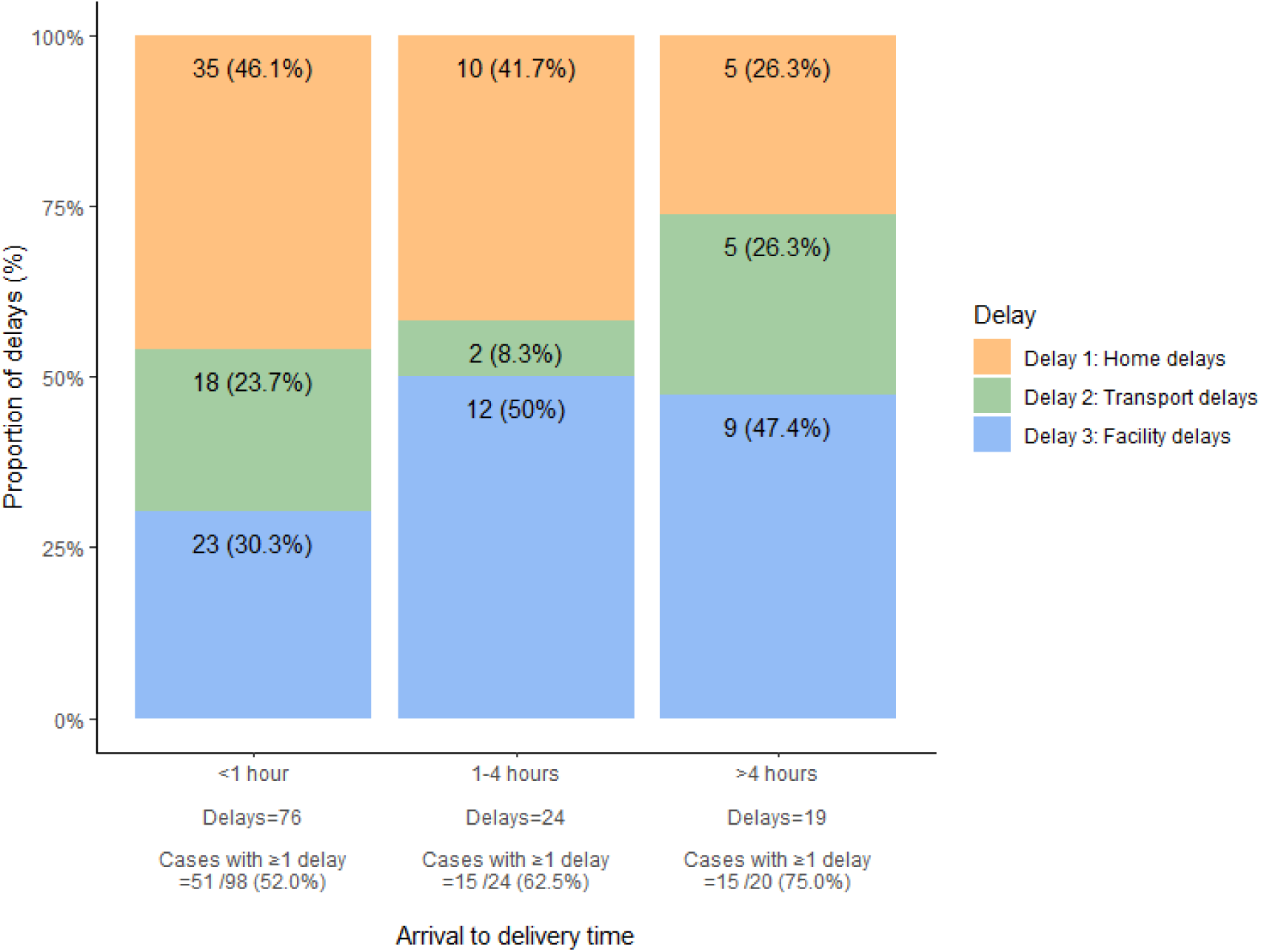
Delays experienced by pregnant women that delivered at the first and nearest facility to where they live, stratified by arrival to delivery time. More than one delay could be reported per case.

## Discussion

To the best of our knowledge, this is the first study to report social autopsy findings for stillbirths and neonatal deaths in Cambodia. Our findings provide crucial insights into healthcare-seeking behaviours and barriers faced by pregnant women experiencing these tragic outcomes in rural Cambodia. Given the slow progress in reducing perinatal mortality rates, such insights can inform policies and programmes aimed at improving stillbirth and neonatal outcomes. Our findings also highlight the value of integrating SA with VA to provide a more comprehensive understanding of the circumstances surrounding these deaths.

Our results demonstrate that pregnant women are responding appropriately to public health advice by seeking facility-based deliveries. The majority (91.1%, 287/315) of women experiencing stillbirths and neonatal deaths reached or attempted to reach a health facility. However, we also found that one-fifth (20.4%, 56/275) of women bypassed their nearest facility, nearly two-thirds (63.7%, 116/182) of women delivered within an hour of arrival to the first facility they attended, and one-third (33.8%, 93/275) left the first facility they attended (transferred out).

The high bypass rate may reflect perceptions and/or previous experiences of care quality at nearby primary facilities.^20,21^ Educating and empowering pregnant women and families on how to negotiate the potentially difficult choice between quickly attending nearer, primary-level care or attending further, higher-level of care could be beneficial.^22^

The high proportion of women delivering within an hour of arrival at a health facility, especially if that facility was a primary facility, is an important finding. Without sufficient time to assess and manage pregnant women, healthcare workers are limited in their ability to deliver the baby safely.^23^ Additionally, referral systems cannot function effectively if there is insufficient time for primary healthcare staff to safely transfer out. Thus, arriving at a facility for delivery is not enough; pregnant women need to arrive with sufficient time to be able to receive quality care. Community and pregnant women’s awareness of the time needed for healthcare staff to delivery their baby safely and provide life-saving care in the event of complications is essential for improving fetal and neonatal health outcomes. Education initiatives should focus on the critical importance of timely arrival for delivery and reinforce a sense of urgency.^22^

Our study also highlights the importance of SA in cause of death reporting.^5^ We show that knowledge of the delivery location alone does not adequately capture the complexities surrounding care-seeking for childbirth. Without SA data, resources might otherwise have been focussed on facilities, where most deliveries (84.1%, 265/315) do indeed occur. However, our data shows that community interventions are still needed to improve demand for facility-based deliveries.

In total, nearly half of all deliveries (49.5%) occurred either at home, enroute, or within one hour of arrival at the first facility attended. These deliveries could be considered to have had no or limited skilled care at birth, and so understanding why this proportion was so high was important. We found that home delays, followed by delays at facilities, particularly regarding transfer, were common. Our findings are in keeping with other studies and underscore the need for tailored interventions across the continuum of care to improve access to skilled care at birth: from improving timely healthcare access in the community (demand) to improving quality of care provision in facilities (supply).^24–26^

To mitigate demand-side issues of home delays, we recommend enhancing education on recognising pregnancy complications and labour signs, and preparing for delivery. ANC appointments provide a great opportunity to educate and advise pregnant women and their families, especially since in our study, almost all pregnant women (97.1%) had at least some antenatal care. Antenatal care should also emphasise practical birth planning, such as knowledge of danger signs and what facility to attend, preparing a facility bag, and establishing home and transport arrangements. Since nearly half of all pregnant women did not save any money in our study, improved financial planning could also be beneficial. With improved birth planning, delays at the time of delivery could be anticipated and avoided, ensuring quicker access to facility-based care.^11^

To mitigate supply-side issues of facility delays, we recommend strengthening of referral systems. Although our study found that women were appropriately being transferred ‘up’ the health system to higher level facilities, deliveries enroute between health facilities suggest weak linkages between health system levels. This highlights the necessity for programmatic and government support to strengthen referral systems and facilitate safer transitions within the healthcare system. Additionally, based on our finding of a high proportion of women delivering very soon after arriving at a facility, it would be prudent to ensure facilities are equipped not just with technical delivery skills but also for rapid response capabilities. High quality of care provision coupled with the ability to handle emergencies has been associated with mortality rate reductions, particularly of intrapartum stillbirths.^6,7,10^

Our recommendations are in keeping with the concept of birth preparedness and complication readiness (BPCR) to reduce delays in receiving skilled care for delivery.^27^ The BPCR interventions were found to be best implemented at multiple health system levels.^26,28^ Similarly, our findings suggest that improving appropriate and timely attendance to facilities (demand) and rapid, quality responses at facilities (supply) may be the key to addressing the disconnect between increasing facility deliveries and falling mortality rates. ^6–9^

## Limitations

The major limitation of this study is the absence of a case-control comparison. Due to resource constraints, it was not feasible to conduct SA on matched controls, such as pregnant women from the nearest village experiencing labour or complications whose infants survived. This limitation hinders our ability to draw any conclusions about the differences in healthcare-seeking behaviours between those who experienced stillbirths or neonatal deaths and those who did not. Nevertheless, this study still provides valuable insights into the social factors associated with perinatal mortality for the first time in Cambodia. Additionally, SA is a resource-intensive method to establish social causes of death, which may limit the scalability.

The reliance on family perspectives for data collection can introduce bias, particularly regarding the recollection of events leading up to the deaths. Respondents were predominantly (98.5%) mothers, whose own condition might have affected their observation of events at the time, particularly if they were experiencing complications themselves. This potential for bias highlights the need for caution when interpreting findings related to the timing and causes of delays in care.

A reliance on the mothers’ perspectives was a limitation of this study.^4^ A lack of the fathers’ perspectives was possibly due to culturally norms as well as SA interviews being conducted during the working day. Furthermore, it was not feasible to review medical records or gather healthcare worker perspectives to triangulate data. Medical records, if available, would likely to have been unreliable. However, healthcare worker perspectives might have been particularly helpful with regards to the facility delays reported by families. Furthermore, respondents, might have presented their experiences through a lens of self-protection or blame, which could distort their accounts of delays or healthcare interactions. Including a broader range of perspectives in future studies could enhance the understanding of factors contributing to stillbirths and neonatal deaths.

While for our population we considered less than one hour from arrival until delivery as insufficient time for adequate healthcare intervention, the optimal arrival time may vary based on individual circumstances, infrastructure, and complications. It is crucial to consider the balance between arrivals that are too late, with insufficient time for healthcare workers to assess and manage the patient, versus arrivals that are too early, which risk pregnant women being turned away and/or adding to already overcrowded facilities. Future research could explore this concept further, gathering stakeholder perspectives that include community members and healthcare providers to determine more context-specific guidance on what is considered a timely arrival for delivery at health facilities. Our recommendation to encourage pregnant women to arrive earlier to facilities should therefore be interpreted with caution.

## Conclusion

Overall, healthcare-seeking behaviour by pregnant women for delivery prior to stillbirths and neonatal deaths was generally appropriate but not timely. Despite high antenatal care uptake and mostly facility-based deliveries, home and facility delays were common and women frequently delivered soon after arrival at facilities. For public health programmes to target improvements in the timeliness of arrival for delivery, more information to better define this would be prudent to avoid any unintended negative consequences. When combined with VA, SA contributes vital upstream information, providing a more comprehensive understanding of the circumstances and causes of death. Thus, targeted strategies that span the continuum of the health system can be implemented to ultimately improve fetal and neonatal health outcomes in low-resource settings.

## Data Availability

All data produced in the present study are available upon reasonable request to the authors

## Abbreviations

SA: social autopsy
VA: verbal autopsy
SBL: Saving Babies’ Lives
NGO: non-governmental organisation
CHW: community health worker
NECHR: National Ethics Committee for Health Research
OxTREC: Oxford Tropical Research Ethics Committee
BPCR: birth preparedness and complication readiness

## Data Availability Statement

The datasets generated and/or analysed during the current study are not publicly available due to ethical and legal reasons, including participant anonymity and privacy, but may be available from the corresponding author on reasonable request. A data access agreement will be put in place prior to data transfer. Instructions and the data application form are available here: https://www.tropmedres.ac/units/moru-bangkok/bioethics-engagement/data-sharing.

## Acknowledgements

The authors wish to thank all community health workers, the Preah Vihear Provincial Health Department, and the Cambodian Ministry of Health for their support of this study. We also thank staff from Angkor Hospital for Children involved with this study, particularly Dr Ngoun Chanpheaktra, Sopheakneary Say, Mara Kik, Lorn Loeuk, Dary Vanna, Khemrouth Cheam, and the fundraising team. Finally, we thank all bereaved women and their families for their participation.

### Appendix 1

Health system structure:

- Within study area (Preah Vihear province):

○ Home
○ Enroute = enroute from home to health facility or between health facilities
○ Government sector:

▪ PrimaryA = Health post (small primary care facility, 1-2 staff, report to health centre)
▪ PrimaryB = Health centre (primary care facility, staffed mostly by midwives and nurses)
▪ SecondaryA = lowest level referral hospital (small secondary care facility – recently upgraded from a health centre just before we started the study, provides assisted delivery but not surgery)
▪ SecondaryB = highest level provincial referral hospital (secondary care facility - provides surgery)
○ Private = private clinic
- Outside study area (Preah Vihear province):

○ Other province Primary / Secondary / Private / NGO (Non-governmental hospital) = as above definitions but not in study area (note all NGO hospitals are tertiary-level hospitals and are located out of province)

### Appendix 2

Appendix 2:
| Traditional practice: Mother | Pregnancy<br>N = 315<sup>1</sup> | Labour<br>N = 315<sup>1</sup> | Postnatal<br>N = 315<sup>1</sup> |
| --- | --- | --- | --- |
| None | 211 (68.3%) | 298 (94.6%) | 102 (32.7%) |
| Tea | 90 (29.1%) | 9 (2.9%) | 188 (60.3%) |
| Other | 3 (1.0%) | 3 (1.0%) | 11 (3.5%) |
| Punley* | 0 (0.0%) | 0 (0.0%) | 7 (2.2%) |
| Unknown | 1 (0.3%) | 0 (0.0%) | 3 (1.0%) |
| Sesame seed | 3 (1.0%) | 0 (0.0%) | 0 (0.0%) |
| Pray | 0 (0.0%) | 3 (1.0%) | 0 (0.0%) |
| Massage | 1 (0.3%) | 1 (0.3%) | 0 (0.0%) |
| Bol** | 0 (0.0%) | 1 (0.3%) | 0 (0.0%) |
| Alcohol | 0 (0.0%) | 0 (0.0%) | 1 (0.3%) |
| Unknown | 6 | 0 | 3 |

<sup>1</sup>n (%)
\* Punley = steam; \*\* Bol = spinning an egg

### Appendix 3

Appendix 3:
| Traditional practice: Hot bed use | Stillbirth<br>N = 127<sup>1</sup> | Neonatal death<br>N = 188<sup>1</sup> | Overall<br>N = 315<sup>1</sup> |
| --- | --- | --- | --- |
| Hot bed use |  |  |  |
| Mother | 76 (60.8%) | 100 (53.2%) | 176 (56.2%) |
| Mother and baby | 0 (0.0%) | 18 (9.6%) | 18 (5.8%) |
| None | 49 (39.2%) | 70 (37.2%) | 119 (38.0%) |
| Unknown | 2 | 0 | 2 |

<sup>1</sup>n (%)

### Appendix 4

Appendix 4:
| Traditional practice: Neonates >12h old | N = 112 |
| --- | --- |
| **General** |  |
| None | 100 (89.3%) |
| Bol* | 4 (3.6%) |
| Punley** | 2 (1.8%) |
| Spitting | 2 (1.8%) |
| Tiger balm | 2 (1.8%) |
| Pray | 1 (0.9%) |
| Other | 1 (0.9%) |
| **Non-medical cord care** |  |
| None | 107 (95.5%) |
| Unknown substance | 2 (1.8%) |
| Salt and pepper | 1 (0.9%) |
| Charcoal and herbs | 1 (0.9%) |
| Saffron | 1 (0.9%) |

\* Bol = spinning an egg; \*\* Punley = steam

### Appendix 5: Facility type and outcomes of pregnancies that delivered at the first facility the pregnant woman attended, by whether they attended or bypassed the nearest facility to them, and further stratified by the time elapsed from arrival to delivery at the facility

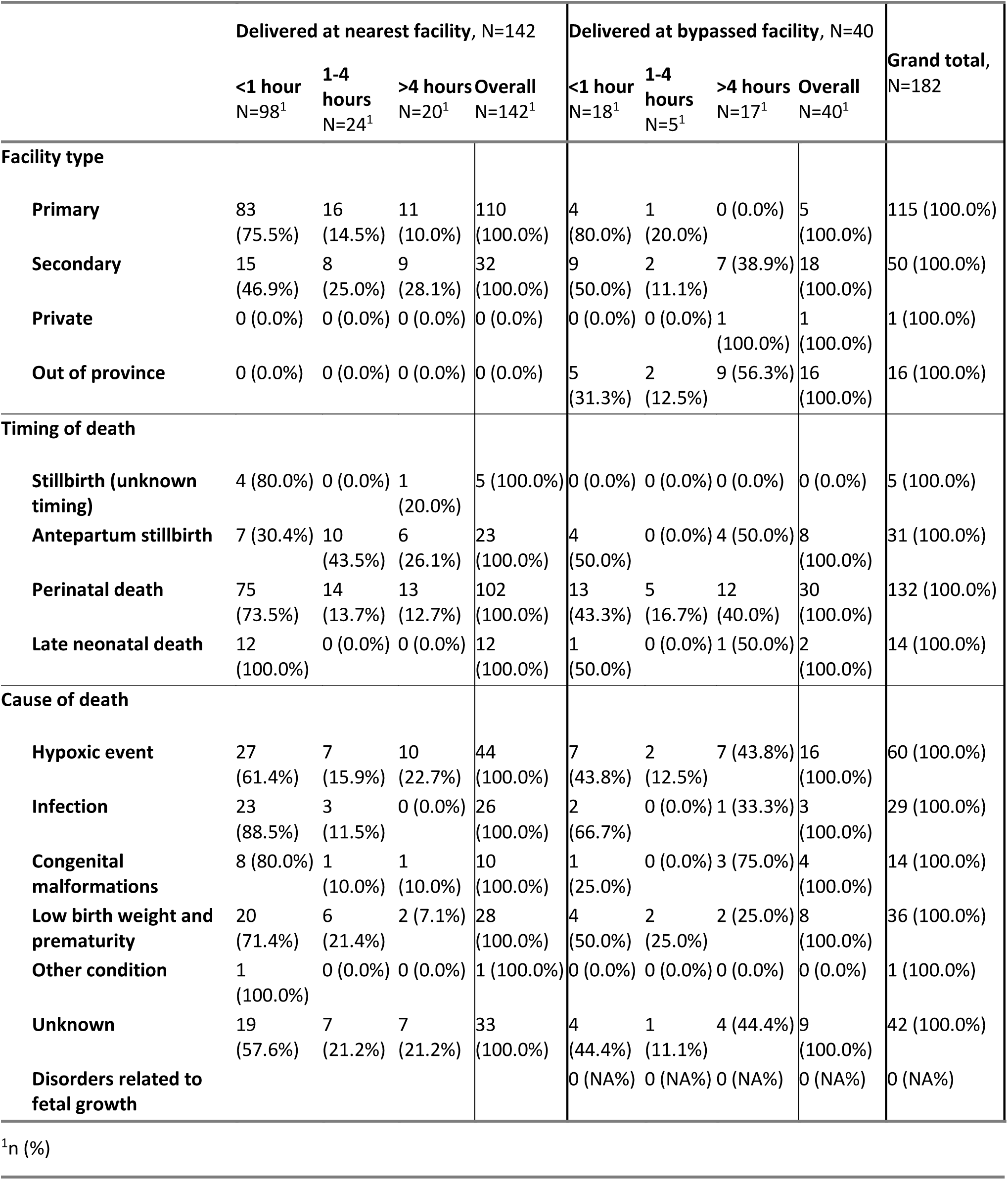

### Appendix 6: Three delays by timing of death

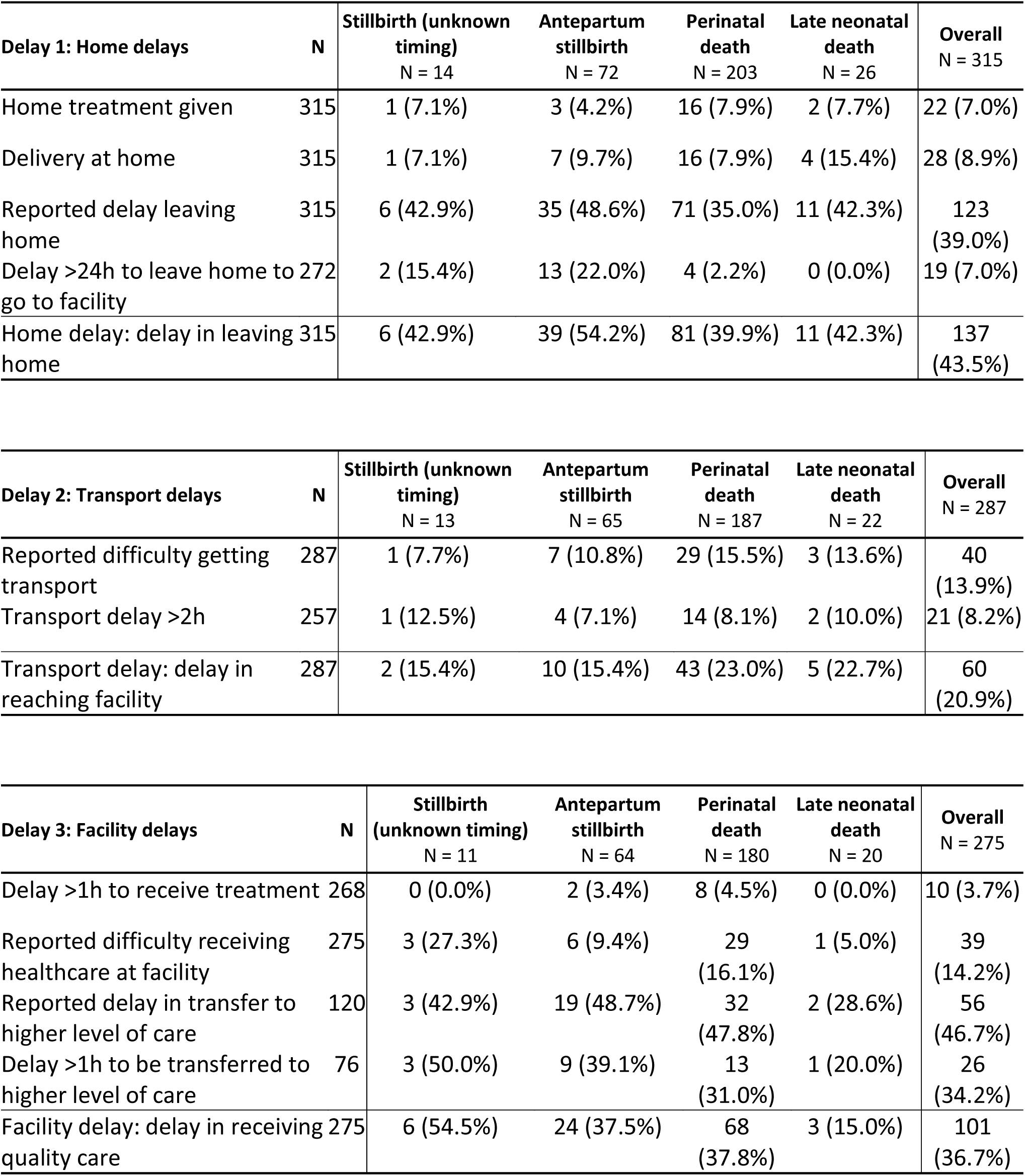

Appendix 6: Three delays by timing of death.
| Delay 1: Home delays | N | Stillbirth (unknown timing)<br>N = 14 | Antepartum stillbirth<br>N = 72 | Perinatal death<br>N = 203 | Late neonatal death<br>N = 26 | Overall<br>N = 315 |
| --- | --- | --- | --- | --- | --- | --- |
| Home treatment given | 315 | 1 (7.1%) | 3 (4.2%) | 16 (7.9%) | 2 (7.7%) | 22 (7.0%) |
| Delivery at home | 315 | 1 (7.1%) | 7 (9.7%) | 16 (7.9%) | 4 (15.4%) | 28 (8.9%) |
| Reported delay leaving home | 315 | 6 (42.9%) | 35 (48.6%) | 71 (35.0%) | 11 (42.3%) | 123 (39.0%) |
| Delay >24h to leave home to go to facility | 272 | 2 (15.4%) | 13 (22.0%) | 4 (2.2%) | 0 (0.0%) | 19 (7.0%) |
| Home delay: delay in leaving home | 315 | 6 (42.9%) | 39 (54.2%) | 81 (39.9%) | 11 (42.3%) | 137 (43.5%) |

| Delay 2: Transport delays | N | Stillbirth (unknown timing)<br>N = 13 | Antepartum stillbirth<br>N = 65 | Perinatal death<br>N = 187 | Late neonatal death<br>N = 22 | Overall<br>N = 287 |
| --- | --- | --- | --- | --- | --- | --- |
| Reported difficulty getting transport | 287 | 1 (7.7%) | 7 (10.8%) | 29 (15.5%) | 3 (13.6%) | 40 (13.9%) |
| Transport delay >2h | 257 | 1 (12.5%) | 4 (7.1%) | 14 (8.1%) | 2 (10.0%) | 21 (8.2%) |
| Transport delay: delay in reaching facility | 287 | 2 (15.4%) | 10 (15.4%) | 43 (23.0%) | 5 (22.7%) | 60 (20.9%) |

| Delay 3: Facility delays | N | Stillbirth (unknown timing)<br>N = 11 | Antepartum stillbirth<br>N = 64 | Perinatal death<br>N = 180 | Late neonatal death<br>N = 20 | Overall<br>N = 275 |
| --- | --- | --- | --- | --- | --- | --- |
| Delay >1h to receive treatment | 268 | 0 (0.0%) | 2 (3.4%) | 8 (4.5%) | 0 (0.0%) | 10 (3.7%) |
| Reported difficulty receiving healthcare at facility | 275 | 3 (27.3%) | 6 (9.4%) | 29 (16.1%) | 1 (5.0%) | 39 (14.2%) |
| Reported delay in transfer to higher level of care | 120 | 3 (42.9%) | 19 (48.7%) | 32 (47.8%) | 2 (28.6%) | 56 (46.7%) |
| Delay >1h to be transferred to higher level of care | 76 | 3 (50.0%) | 9 (39.1%) | 13 (31.0%) | 1 (20.0%) | 26 (34.2%) |
| Facility delay: delay in receiving quality care | 275 | 6 (54.5%) | 24 (37.5%) | 68 (37.8%) | 3 (15.0%) | 101 (36.7%) |

### Appendix 7: Number and percent of delays, by timing of death, experienced by pregnant women prior to the delivery of a stillbirth or neonatal death. More than one delay could be reported per case

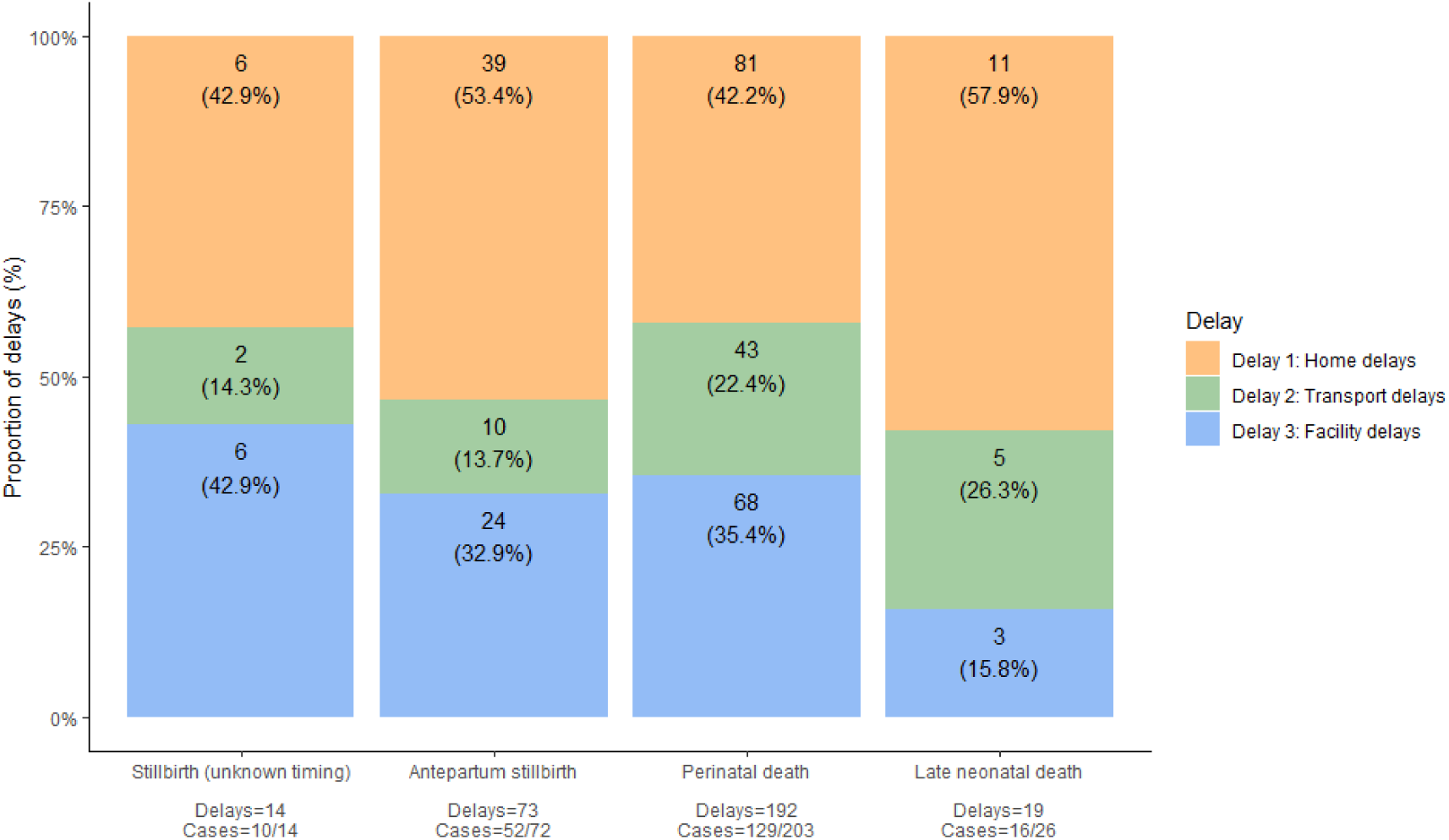

### Appendix 8: Number and percent of delays, for the top four known medical causes of death, experienced by pregnant women prior to the delivery of a stillbirth or neonatal death. More than one delay could be reported per case

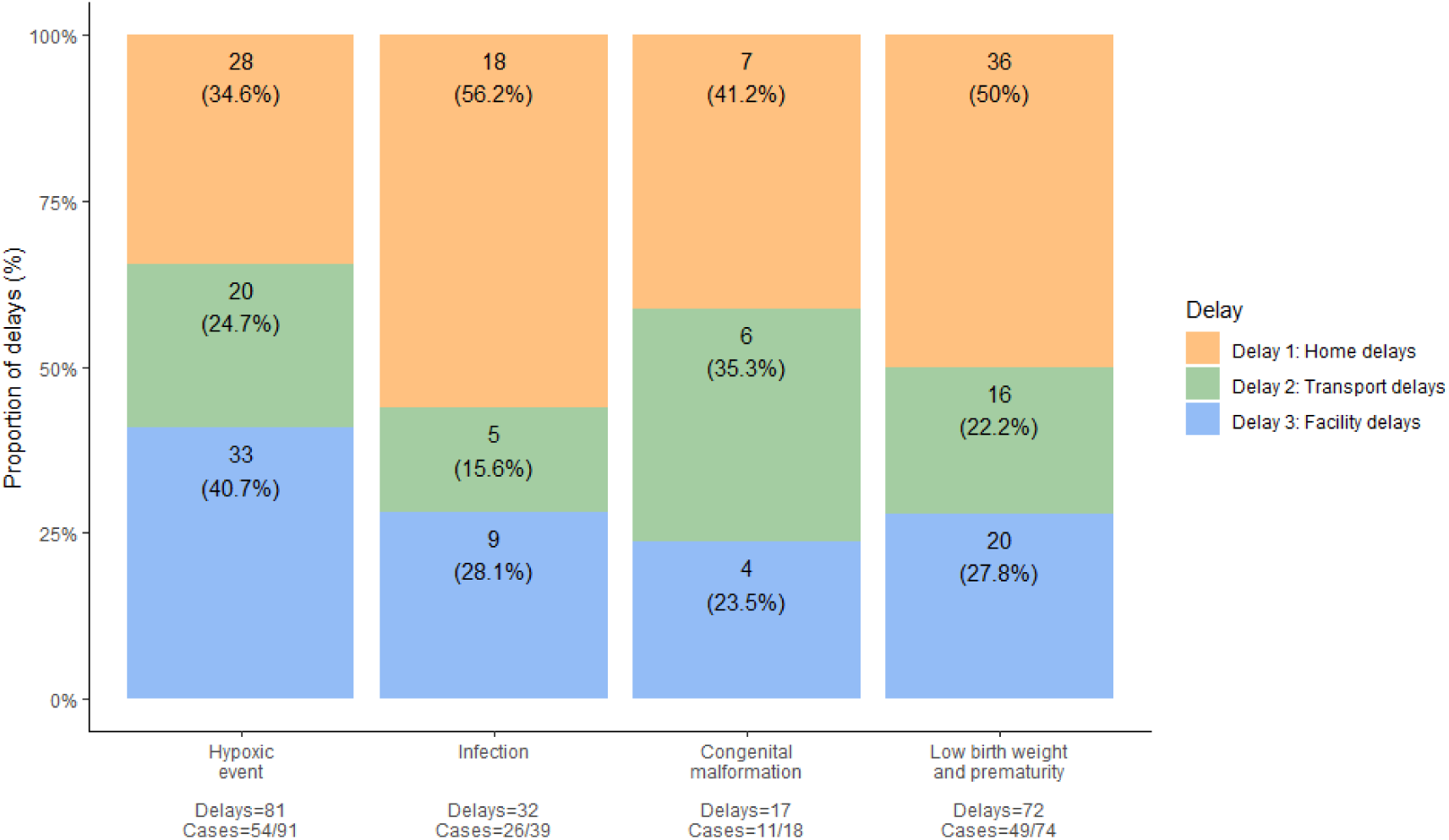

### Appendix 9: Reported reasons for delays leaving home and delays in transfer

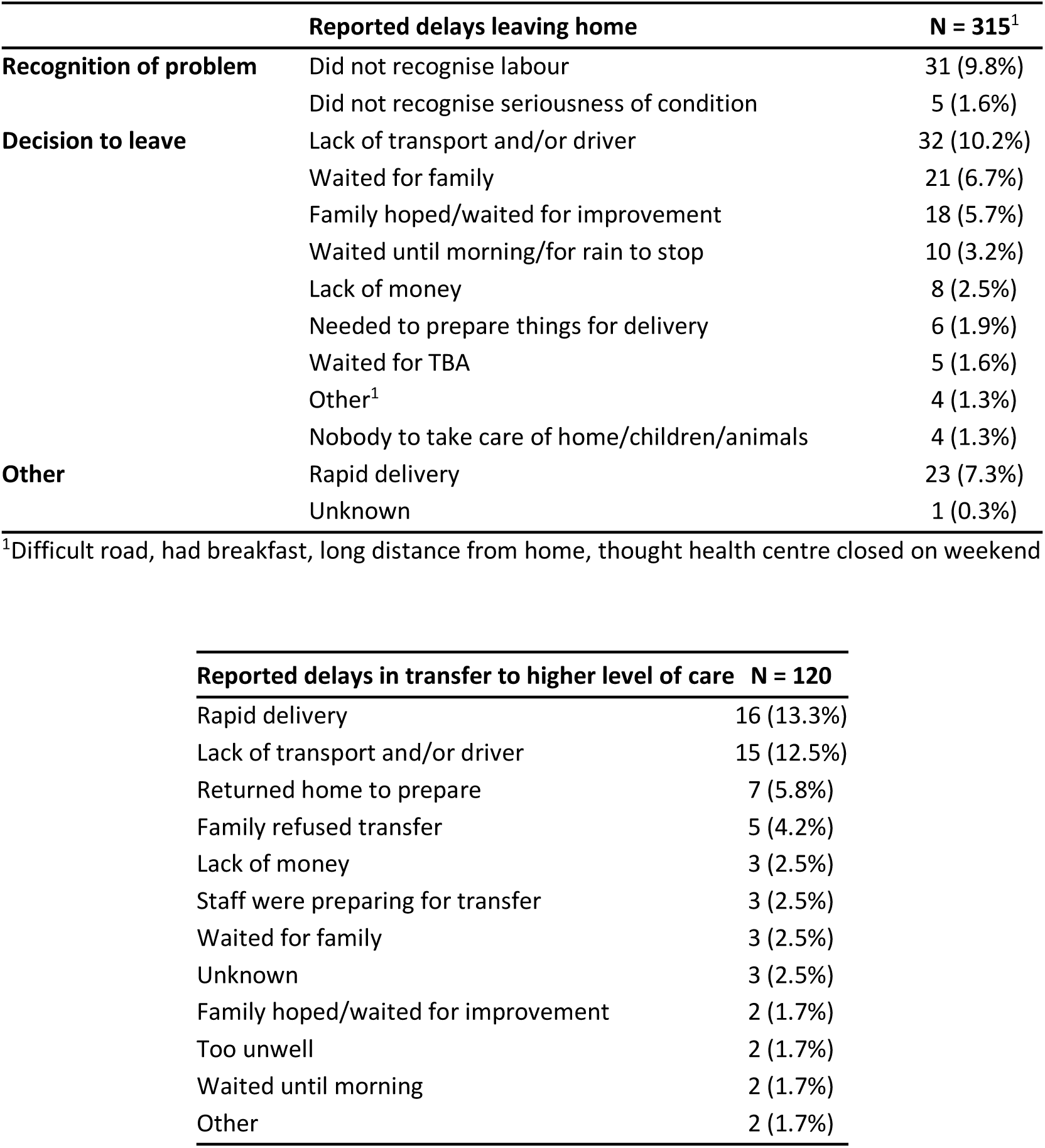

